# First trimester serum Cystatin C level is a predictor of Preeclampsia: a prospective study [Correlation between Shrunken pore syndrome and Preeclampsia]

**DOI:** 10.64898/2025.12.06.25341768

**Authors:** Pooja Prakash Prabhu, Mahesh Eshwarappa, Krishnamoorthy, B.K Sujani, Prachi, Gurudev K.C

## Abstract

**Introduction:** The pathogenesis of Preeclampsia (PE) remains enigmatic. A new concept of Shrunken Pore Syndrome (SPS) has been proposed, where in there is shrinking of pore size of the glomerulus. The ratio eGFR_cystatin C_ / eGFR_creatinine_ < 0.6 is considered as SPS. Few studies show that SPS exists in pregnant ladies in the first trimester. A novel biomarker which can predict preeclampsia in the first trimester is the need of the hour. Hence this study was done.

**Method:** Cystatin C and creatinine levels were estimated during 11 to 13 weeks in all pregnant ladies. The GFR was calculated based on creatinine and cystatin C, and were assessed for SPS. Blood pressure was monitored and was managed appropriately, and statistically analysed.

**Results:** 120 pregnant ladies were enrolled in our study. Out of 120, 91 ladies had SPS. 17 ladies developed preeclampsia during the follow up. The cystatin C levels were significantly higher in those who developed preeclampsia. A first trimester serum cystatin C value of 1.24mg/dl has shown to predict the occurrence of preeclampsia, with sensitivity of 94% and specificity of 74.8%. All 17 patients who had preeclampsia, had an evidence of SPS. First trimester serum cystatin C value was higher among those subsequently developed preeclampsia.

**Discussion:** There is a positive correlation between maternal first trimester serum cystatin C levels and development of preeclampsia. Assessment of Serum Cystatin C levels in the first trimester is a novel, cost effective biomarker to determine the risk of preeclampsia and improve the maternal, fetal and renal prognosis.

## INTRODUCTION

Preeclampsia is a common complication noted in pregnancy, affecting 3-8% of all pregnancies (1). It is associated with increased risk of maternal and fetal mortality and morbidity. The exact pathogenesis of this disorder remains enigmatic. Various biomarkers and inflammatory mediators have been proposed to be involved in the process and several studies have been done across the globe to identify the molecular pathway affected. Renal damage in preeclampsia is one of the key components of the pathophysiological process in preeclampsia. Serum creatinine and uric acid levels are routinely used as markers of renal damage. Recently, serum Cystatin C levels have been shown to be a better marker of renal damage than serum creatinine in Preeclampsia.

A new concept of Shrunken Pore Syndrome (SPS) has been proposed by A. Grubb, where in it has been noted that there is a pronounced shrinking of pore size of the glomerulus during preeclampsia (2). This has also been seen in 5% of healthy men and non-pregnant women. Due to shrinkage of the pore size, serum creatinine being a smaller molecule gets filtered away, but cystatin C (MW-133 kDa) is retained in circulation. Thus the serum levels of cystatin C is higher than serum creatinine levels and hence the eGFR_cystatin C_ will be lesser than eGFR_creatinine_. The ratio eGFR_cystatin C_ / eGFR_creatinine_ < 0.6-0.7 has been used to diagnose SPS. This raised the question whether those ladies who have SPS in the first trimester are at an increased risk of preeclampsia? There are very few studies which have shown that serum cystatin C could be a useful marker than serum creatinine in the first trimester to predict preeclampsia. Some studies have shown that SPS did exist in pregnant ladies in the first trimester who later on went to develop preeclampsia.

A novel biomarker which can predict preeclampsia in the first trimester helps to be more vigilant, enable us to institute risk modifying therapies at the earliest and improve maternal, neonatal and renal prognosis. Thus the objectives of the study were to determine the serum creatinine and serum cystatin c levels among all the pregnant ladies presenting in the first trimester, to calculate eGFR_Cystatin C_ and eGFR_creatinine,_ and evaluate for shrunken pore syndrome, And to look for any association between first trimester serum cystatin c levels with the development of pre-eclampsia.

## METHOD

This study was done in collaboration with the three departments of Nephrology, Obstetrics and Gynecology, and Biochemistry. It was a prospective analytical study done between January 2022-December 2022. All pregnant ladies in the first trimester presenting to the department of OBG during the study period were enrolled into the study. All pregnant ladies > 18 years of age, in their first trimester (11-13 weeks of gestation) were included in the study. Those who had co-morbidities like Type 2 Diabetes mellitus, chronic hypertension, Hypothyroidism, those who had conceived with Assisted Reproductive Technology, those who were on anticoagulants were excluded from the study. The patient clinical and obstetric details were entered into the proforma. After obtaining informed consent, blood samples were collected for estimation of serum cystatin c and serum creatinine levels. A baseline urine routine was done in the first trimester. Serum creatinine was analyzed by Jaffe’s method, traceable to IDMS assigned NIST certified reference material. Serum Cystatin C was assayed using turbidometric method that was traceable to IFCC and IRMM certified reference material.

Patients were regularly followed up every month during their antenatal visits to determine the onset of preeclampsia at the earliest. Those patients who develop preeclampsia –as per definition, were further investigated. These patients were monitored till delivery and renal functions were monitored on need basis. The fetal growth was monitored by routine antenatal scans. The decision to terminate the pregnancy was based on severity of preeclampsia and obstetrician discretion. The maternal and fetal outcomes were documented and entered in the proforma.

### Statistical analysis

Data was summarized using Mean +/-standard deviation. Categorical variables were compared with □^2^ test. Continuous variables were compared using t-test or Mann Whitney U test. Multivariate regression analysis were performed to assess the association between cystatin C levels and preeclampsia. All data analysis was done using SPSS software version 28.

## RESULTS

One hundred and twenty pregnant ladies in their first trimester were enrolled in our study. 10% were aged more than 30 years. 56 were primi gravida. 10 out of 120 ladies had history of preeclampsia in their previous pregnancies, of which 9 had maternal history of preeclampsia. The mean serum creatinine in our population was 0.46mg/dl (SD-0.09) with average eGFR_creatinine_ being 142ml/min/ 1.73m^2^ BSA. The mean cystatin-C level was 1.1mg/dl (SD-0.26), with average eGFR_cystatin C_ being 75ml/min/1.73m^2^BSA. There is a statistically significant correlation between the GFRs in pregnancy, calculated by eGFR_creatinine_, eGFR_cystatinC_ and eGFR _(creatinine + cystatin C)_(fig 1). The Area under the Curve (AOC) for cystatin C is 0.873, which was statistically significant (p<0.0001) (fig 2).

**Fig 1:**
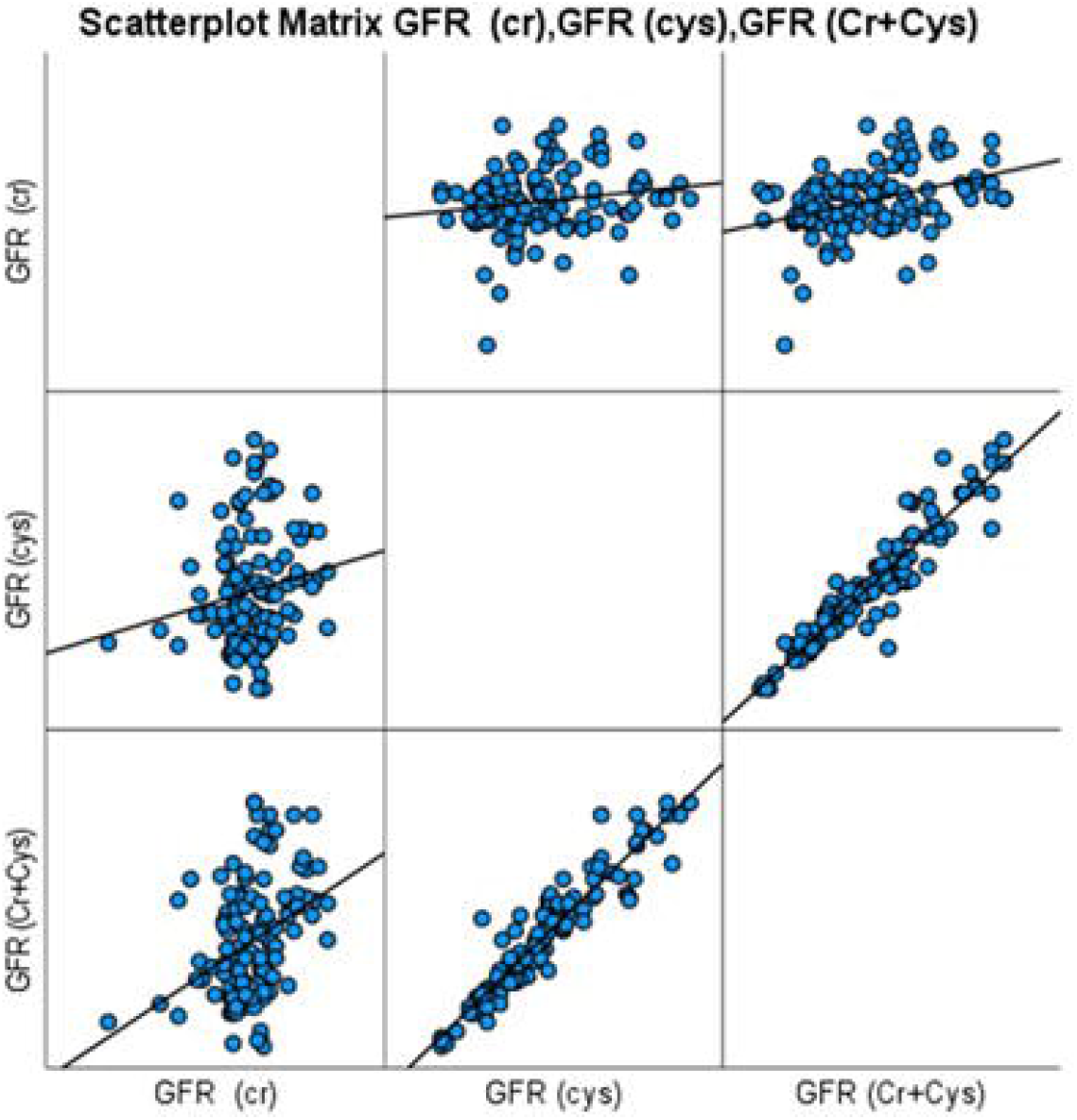
Scatter Plot Matrix showing correlation between the GFRs calculated by creatinine, cystatin Cand creatinine + cystatin C

**Fig 2:**
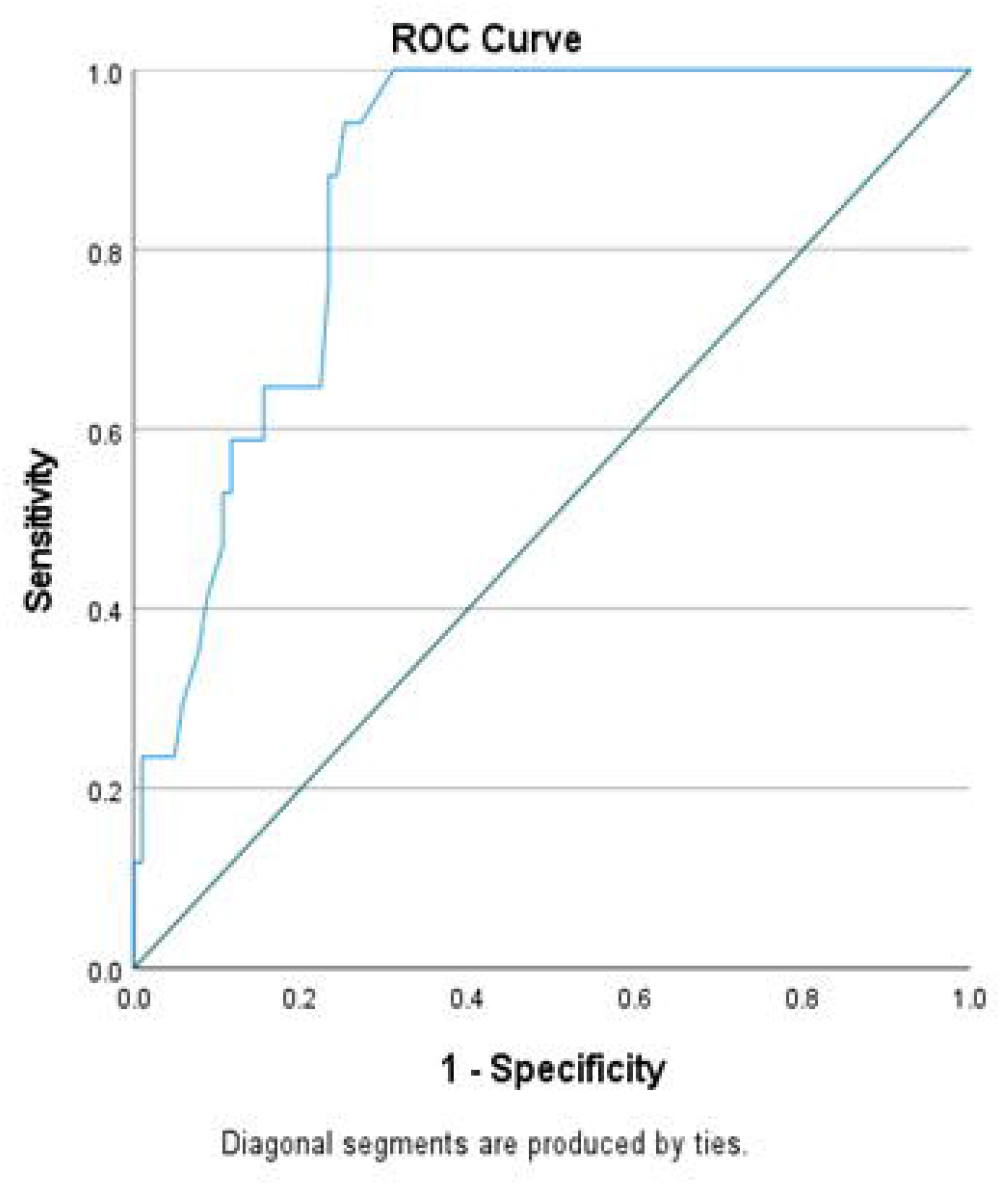
ROCcurve for cystatin C-0.86 (p-0.000)

Seventeen pregnant ladies went on to develop preeclampsia during the follow up. The cystatin C levels in the first trimester were significantly higher in those who developed preeclampsia, compared to those who had normal blood pressure (table-1). A serum cystatin C value of 1.24mg/dl in the first trimester has shown to predict the occurrence of preeclampsia, with a Youdon Index sensitivity of 94% and specificity of 74.8%.

**Table 1:** Correlation between first trimester serum creatinine and cystatin C values and development of Preeclampsia.

|  |  |
| --- | --- |
|  | **Preeclampsia (PE)** |

|  | Yes |  | No |  | p-value |
| --- | --- | --- | --- | --- | --- |
|  | Mean | SD | Mean | SD |  |
| Creatinine | 0.50 mg/dl | 0.11 | 0.46 mg/dl | 0.08 | 0.08 |
| Cystatin C | 1.45 mg/dl | 0.216 | 1.10 mg/dl | 0.235 | **<0.001** |
| eGFR (cr) | 138 ml/min | 14.714 | 142 ml/min | 10.045 | 0.1 |
| eGFR (cys) | 52 ml/min | 9.65 | 78 ml/min | 22 | **<0.001** |

Among the 120 pregnant ladies, 91 (75.8%) had evidence of Shrunken Pore Syndrome (SPS). All 17 patients who had PE had evidence of SPS, which was statistically significant. The serum cystatin C values was significantly higher among those who had SPS and subsequently developed preeclampsia (table-2).

**Table 2:** Correlation between the presence of shrunken pore syndrome (SPS) and development of preeclampsia in pregnancy.

|  |  | Preeclampsia (PE) |  |  |
| --- | --- | --- | --- | --- |
|  |  | Yes | No | Total |
| SPS | No | 0 (0.0%) | 29 (100%) | 29 |
|  | Yes | 17 (18%) | 74 (81%) | 91 |

Among those pregnant ladies who had preeclampsia, 7 had AKI, 2 abruptio placenta, 6 had HELLP syndrome, 1 had Eclampsia. Out of the 7 who had AKI, 6 showed complete recovery after delivery, 1 patient had persistent renal dysfunction (creatinine-1.4mg/dl) at 3 months of follow up. Fetal morbidity was also noted among the babies delivered in the preeclamptic mothers. 4 were preterm and 5 were SGA (small for gestational age) requiring NICU care. There was one fetal demise.

## DISCUSSION

Renal damage in PE is one of the key components of the pathophysiological process and among the most significant event which leads to the endothelial dysfunction and decline in glomerular filtration rate (GFR)^[1]^. Thus, the most commonly used markers in monitoring renal dysfunction in pregnancy with PE are serum concentration of creatinine and uric acid^[3]^. Several authors have also proposed serum Cystatin C levels as a possible alternative marker of renal dysfunction^[4-6]^. Cystatin C is a 13-kDa protein, a member of the cysteine proteinase inhibitors family, which is produced at a constant rate in all nucleated cells, freely filtered across the glomerular membrane, and finally metabolized in the proximal tubules^[1]^. Various studies have shown that cystatin C has superior diagnostic accuracy as a marker of glomerular endotheliosis or renal involvement in preeclampsia compared to creatinine^[7-8]^.

There is a reduction in the serum creatinine level as the pregnancy progresses. A study by Agampodi et.al ^[9]^ has shown that the average serum creatinine value was 84.7% and 76.4% of the non-pregnant group, respectively, in the first and second trimesters. In our study, we enrolled 120 pregnant ladies in the first trimester and evaluated the renal functions in them. We found that there was a significant difference between the mean serum creatinine and mean serum cystatin C levels in the first trimester. There was a statistically significant difference between the GFR calculated based on creatinine and cystatin C levels. We lack regional data on average kidney function in the first trimester among our pregnant ladies. To the best of our knowledge, this is the first study from South India to have systematically evaluated the pregnant ladies for any renal dysfunction in their first trimester.

Grubb et.al has done studies on the plasma levels of cystatin C, beta 2 microglobulin and beta trace proteins in pre eclampsia, and has showed that the levels of these proteins were higher indicating a pronounced shrinking of pore size, which is a feature of pre eclampsia ^[2]^. Thus the eGFR_Cystatin C_ will be lesser than eGFR_Creatinine_ among preeclampsia. This phenomenon has also been noted among healthy males and non pregnant females^[2]^. Various studies have been done to establish a reliable reference values for eGFR_cystatin C_ / eGFR_creatinine_ ratio in order to diagnose Shrunken Pore Syndrome. In most studies of SPS, eGFR_cystatin C_ / eGFR_creatinine_ ratio of 0.6 - 0.7 has been used and it has high specificity and good sensitivity for predicting 3-6 year mortality among general population ^[10,11]^.

There are very few studies which have assessed for the presence of SPS in the first trimester of pregnancy. A study done by Campos et.al ^[12]^, where in 74 pregnant ladies were evaluated in their first trimester for serum cystatin C and serum creatinine levels. Eight of them had SPS. In our study, though none of the pregnant ladies had elevated serum creatinine (>0.8mg/dl) in the first trimester, 109 out of 120 had a higher serum cystatin C levels (>0.85mg/dl) in the first trimester. There was a very high prevalence of SPS (75%) in our cohort. This could be probably explained by the very low serum creatinine levels among our female population as most of them were from rural areas, belonging to low socio economic strata and with a lower muscle mass. In our study, seventeen pregnant ladies went on to develop preeclampsia during the follow up. All 17 patients had the evidence of SPS. The serum cystatin C values were significantly higher among those who had SPS and subsequently developed preeclampsia. A study by Risch et.al^[13]^ showed that women who later developed PE had significantly higher levels of Cystatin C in their first trimester. Among the PE cases, 5 of them had a higher value of cystatin C and eGFR_cystatin C_ / eGFR_creatinine_ was significantly lower. Thus they concluded that a serum cystatin C value >0.85 mg/L in the first trimester despite a normal serum creatinine, showed the evidence of altered glomerular filtration quality, suggesting the presence of glomerular endotheliosis. Similarly, a study done by Thilaganathan et.al ^[14]^ also demonstrated that raised maternal first trimester serum cystatin c as an early marker of pre eclampsia. In our study too, we concluded that a serum cystatin C value > 1.24mg/dl in the first trimester predicted the occurrence of preeclampsia, with a Youdon Index sensitivity of 94% and specificity of 74.8%.

## CONCLUSION

Pre eclampsia is an increasingly prevalent pregnancy complication that develops in the second half of gestation. It is associated with maternal and fetal mortality and morbidity. There are various biomarkers which have been studied in pre eclamptic women, but there is no highly accurate method for determining the general risk of PE, especially a few weeks before its onset. Serum cystatin C level seems to be a useful first trimester biomarker for prediction of preeclampsia. Determining the risk of development of PE in the first trimester will help in early treatment of these women with low dose aspirin, which has shown to substantially reduce the incidence of PE and some of its complications.

## Data Availability

All data produced in the present work are contained in the manuscript

## Authors’ Contributions

Authors Contributions (Example)

**Pooja Prakash Prabhu**: Conceptualization, writing original draft, data curation

**Mahesh Eswarappa**: Project administration, Resources

**Krishnamoorty:** Formal analysis, Investigation, formal analysis

**Sujani B.K:** Resources, Visualization

**Prachi Bajaj:** Investigation

**Gurudev K.C:** Funding acquisition and supervision

## Data Availability

**YES**

## Definitions

### 1. Preeclampsia (Diagnostic Criteria by American College of Obstetricians)

a. **Blood pressure**
  - Systolic BP > 140mm of Hg or higher, Diastolic BP of 90 mm of Hg or higher on two occasions atleast 4 hours apart after 20 weeks of gestation in a woman with a previously normal blood pressure.
  - Systolic BP of 160 mm of Hg or higher, or Diastolic BP of 110 mm of Hg or higher. AND
b. **Proteinuria**
  - 300 mg per 24 hour urine collection
  - Urine protein creatinine ratio >0.3
  - Dipstick reading of 1+

### Or in the absence of proteinuria, a new onset hypertension with new onset any of the following

1. Thrombocytopenia - Platelet counts < 1lakh/ml
2. Impaired liver function – Elevated liver enzymes (twice the normal concentration) or persistent right upper quadrant pain
3. Renal insufficiency – Serum creatinine values > 1.1 mg/dl or Doubling of serum creatinine levels
4. Pulmonary oedema
5. Cerebral/ Visual symptoms.

### 2. Shrunken Pore Syndrome

A ratio of eGFR_cystatin C_ / eGFR_creatinine_ <0.6 is considered as suggested by Grubb et.al [13]

### 3. Glomerular filtration rate

The National Kidney Foundation has recommended to use CKD EPI equation to determine GFR in adults, developed in 2009 by Chronic Kidney Disease –Epidemiology Collaboration (CKD-EPI)

### eGFR_creatinine_ = 141 * min (S_cr_ /k, 1)^α^ * max (S_cr_ /k, 1)^-1.209^ * 0.993^age^ *

#### 1.018 (if female) * 1.159 (if black) eGFR= Estimated

GFR (ml/min/1.73 sq.m BSA)

Scr= Standardized serum creatinine (in mg/dl)

K= 0.7 (females) or 0.9 (males)

α = -0.329 (females) or -0.411 (males)

min= indicates the minimum of S_cr_ / k or 1

max= indicates the maximum of S_cr_ / k or 1

Age = in years

### eGFR_cystatin C_ = 133 * min (S_cys_ /0.8, 1)^-0.499^ * max (S_cys_ /0.8, 1)^-1.328^ * 0.996^age^ * 0.932 (if female)

S_cys_ (Standardized serum cystatin c) in mg/dl

Min= indicates a minimum of S_cys_ / 0.8 or 1

Max = indicates the maximum of S_cys_ / 0.8 or 1

Age = in years

